# Antimicrobial peptides and other potential biomarkers of critical illness in Sars-CoV-2 patients with acute kidney injury

**DOI:** 10.1101/2023.07.08.23292389

**Authors:** Lucas Ferreira Theotonio dos Santos, Hermes Vieira Barbeiro, Denise Frediani Barbeiro, Heraldo Possolo de Souza, Fabiano Pinheiro da Silva

## Abstract

Antimicrobial peptides (AMPs) are a complex network of 10-100 amino acid sequence molecules, widely distributed in Nature. Even though more than 300 AMPs have been described in mammals, cathelicidins and defensins remain the most investigated to date.

Some publications examined the role of AMPs in COVID-19, but the findings are preliminary and *in vivo* studies are still lacking. Here, we report the plasma levels of five AMPs (LL-37, α-defensin 1, α-defensin 3, β-defensin 1 and β-defensin 3) and five cytokines (tumor necrosis factor-α, interleukin-1β, interleukin-6, interleukin-10, interferon-γ and monocyte chemoattractant protein-1), in 15 healthy volunteers, 36 COVID-19 patients without Acute Kidney Injury (AKI) and 17 COVID-19 patients with AKI, since AKI is a well-known marker of worse prognosis in Sars-CoV-2 infections.

We found increased levels of α-defensin 1, α-defensin 3 and β-defensin 3, but not LL-37 or β-defensin 3, in our COVID-19 population, when compared with the healthy controls, in conjunction with higher levels of interleukin-6, interleukin-10, interferon-γ and monocyte chemoattractant protein-1, putting in evidence that these AMPs and cytokines may have an important role in the systemic inflammatory response and tissue damage that characterizes severe COVID-19.

**Graphic Abstract:** 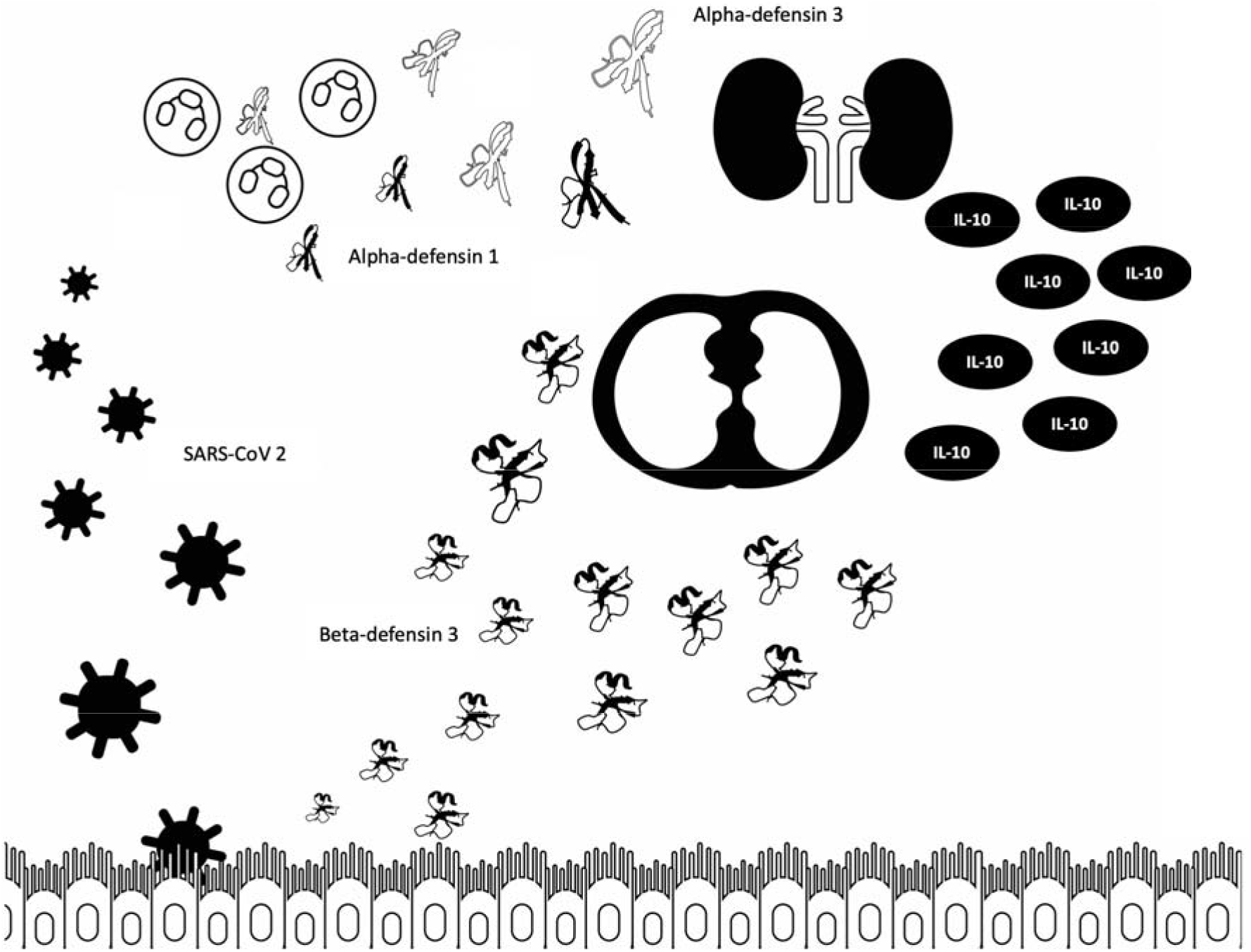

## Introduction

Antimicrobial peptides (AMPs), also known as host defense peptides (HDP), comprise a broad number of molecules, present in both eukaryotes and prokaryotes. Initially investigated for their direct antimicrobial properties, further studies are increasingly putting in evidence that they are major regulators of the immune response [1]. In higher organisms, cathelicidins, α- and β-defensins are the most widely investigated AMPs.

Humans and rodents contain only one cathelicidin family member, produced by immune and epithelial cells and named, respectively, LL-37 and CRAMP (cathelin-related antimicrobial peptide). Defensins, by the other hand, are divided in three main classes in mammals: the α-, β- and θ-defensins. θ-defensins, however, are only found in Old World monkeys. Human α-defensins 1-4 are mainly present in neutrophils, whereas human β-defensins, in a similar way to cathelicidins, are produced by several types of immune and epithelial cells.

COVID-19 (Coronavirus Disease 2019) is an infectious disease caused by severe acute respiratory syndrome coronavirus 2 (Sars-CoV-2). It was first described in China in 2019 and gained global attention, since it rapidly became a devastating pandemic. Worldwide, around 3,5 million people already died because of this disease, which has a diverse clinical course, but generally includes signs and symptoms of systemic inflammation. Even though the lung is the most affected organ, Sars-CoV-2 severe infections frequently lead to Acute Kidney Injury (AKI) [2, 3]. The mechanisms of kidney damage still need to be fully elucidated, but a combination of direct viral damage to the renal cells, systemic inflammation and endothelial dysfunction have been implicated [4, 5]. AKI is more common in hospitalized COVID-19 patients, being a marker of worst prognosis and increased mortality [6, 7].

Antimicrobial peptides have been poorly investigated in COVID-19. A recent study demonstrated *in vitro* that LL-37 not only binds to Sars-CoV-2 spike 1 protein, but also cloaks the ligand-binding domain of angiotensin-converting enzyme 2 (ACE2), protecting against virus entry into the cell[8]. In addition, there are reports that α-defensins 2 and 5 also protect epithelial cells *in vitro* from Sars-CoV-2 invasion, using an analogous mechanism [9, 10]. However, for a better comprehension of the role of AMPs in severe COVID-19, *in vivo* studies are still lacking. Abdeen et al. showed that α-defensins plasma levels are elevated in COVID-19 patients, but they don’t specify which α-defensin was measured [11]. Several studies that investigated the role of vitamin D in Sars-CoV-2 infections, moreover, don’t comment the importance of LL-37 in this scenario [12, 13], even though vitamin D is a potent inducer of cathelicidin gene expression [14].

Here, we report the plasma levels of 5 antimicrobial peptides (LL-37, α-defensin 1, α-defensin 3, β-defensin 1 and β-defensin 3) in 15 healthy subjects, 36 COVID-19 patients without AKI and 17 COVID-19 patients with AKI, in conjunction with the measurement of several cytokines (tumor necrosis factor-α, interleukin-1β, interleukin-6, interleukin-10, interferon-γ and monocyte chemoattractant protein-1). These AMPs were selected, because they have been implicated in several other systemic inflammatory states [15-17]. HBD-1 is expressed in kidney epithelial cells, among others, and HBD-3 is produced by lung epithelial cells [18-20]. In situ hybridization localized the HBD-1 mRNA in the epithelial layers of the loops of Henle, distal tubules, and the collecting ducts of the kidney [21]. Given that finding, one may suppose that HBD-1 is a potential biomarker of tubular damage. Overall, all these antimicrobial peptides are potential biomarkers of disease activity, organ damage and risk of death.

## Methods

### Selection of patients and samples collection

This study was approved by the Ethics Committee of our hospital (protocol # 30417520.0.0000.0068). An informed consent form was signed by the responsible family member, for inclusion of the patient in our study.

A total of 53 patients were included, out of a database 1498, after applying exclusion criteria (Figure 1). They were divided in two groups, according to the KDIGO criteria [22]: patients without AKI (COVID-19 group, n=36) and patients with AKI (COVID-19 AKI group, n=17). The admission creatinine was used as baseline. The control group consisted of 15 healthy volunteers.

**Figure 1.**
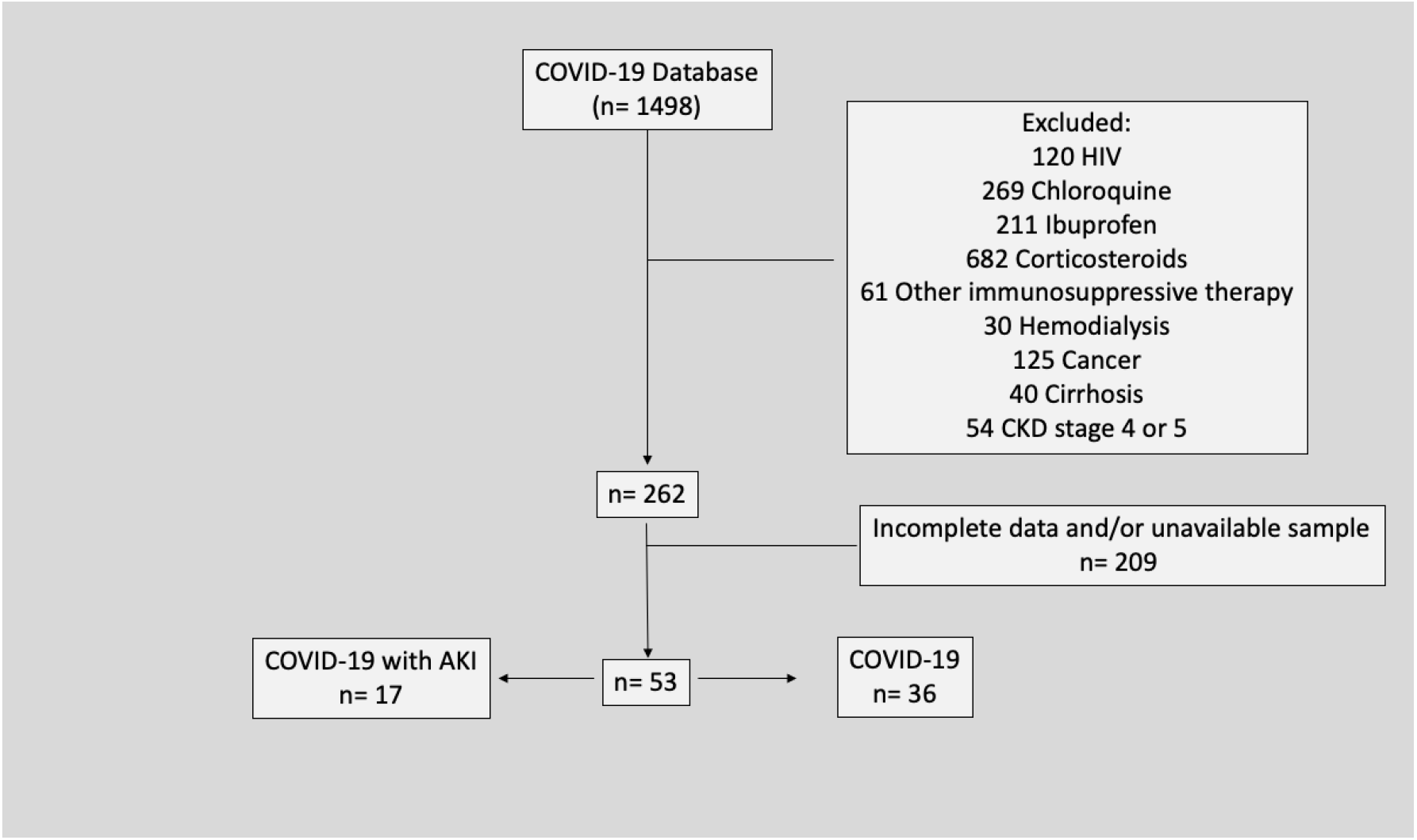
Patients selection

Patients aged 50 years and over, with COVID-19 diagnosis confirmed by real-time polymerase chain reaction (RT-PCR) and at least 2 serum creatinine values during in-hospital stay were included. Healthy individuals aged 50 years and over, without symptoms of COVID-19 in the prior 30 days and without exclusion criteria were selected.

Patients with diagnosis of cancer, chronic liver disease, HIV/AIDS, chronic kidney disease and in use of immunosuppressive agents were excluded.

Blood samples were collected within 24h of hospital admission, plasma was immediately separated by centrifugation and stored at -80°C for further analyses.

### Antimicrobial peptides and cytokines measurements

α-defensins 1 and 3, β-defensins 1 and 3 and LL-37 plasma levels were measured using the ELISA technique (MyBioSource, San Diego, CA, United States). Tumor necrosis factor-α (TNF-α), interleukin-1β (IL-1 β), interleukin-6 (IL-6), interleukin-10 (IL-10), interferon-γ (IFN-γ) and monocyte chemoattractant protein-1 (MCP-1) plasma levels were measured using the magnetic bead immunoassay Milliplex® and the MAGPIX® System (MilliporeSigma, Darmstadt, Germany).

### Statistical analysis

Continuous clinical characteristics and laboratory parameters were analyzed using the Kruskal-Wallis test or the Mann-Whitney test, as appropriate. The Chi-Square test of independence was used for categorical clinical values. Results are shown as medians and interquartile ranges for the continuous variables or medians and percentages for the categorical values. Plasma measurements were analyzed using the Kruskal-Wallis test, followed by the Mann-Whitney U test for *post hoc* analysis. ROC curve analysis was performed to test the of cytokines and antimicrobial peptides to discriminates those with AKI from those with not. Correlation was measured through Pearson correlation test, and a coefficient (r) of 0.5 or greater, or -0.5 or less, was considered significant. All analyses were performed using R statistical software (www.r-project.org). A p-value ≤ 0.05 was considered significant.

## Results and Discussion

### In-hospital routine laboratory parameters were poor indicators of disease severity in our COVID-19 population

No difference could be detected regarding age, gender or presence of comorbidities among the study groups, demonstrating that the individuals share a very similar profile (Table 1). The COVID-19 AKI group, however, exhibited longer hospital stay, higher rate of Intensive Care Unit (ICU) admission, more days in mechanical ventilation and higher mortality (Table 1), confirming that COVID-19 patients with AKI have worse prognosis. Table 2 exhibits the values of several laboratory parameters obtained from our COVID-19 patients’ blood, within 24 hours hospital admission. Blood gases, urea, creatinine, electrolytes, number of blood cells, C-reactive protein and D-dimer values were investigated. Interestingly, we could detect no statistical differences, when the COVID-19 and the COVID-19 AKI groups were compared, putting in evidence that these parameters are poor indicators of inflammatory state, disease severity or risk of death in this situation. A recent study from our university, however, also investigated several routine laboratory parameters in COVID-19 patients and found interesting differences, when patients are grouped by gender or age [23].

**Table 1.** Clinical characteristics of the healthy individuals (n = 15) and patients included in our study (COVID-19 group = 36 patients; COVID-19 AKI group = 17 patients). Mann-Whitney test was used to compare symptoms duration, days in mechanical ventilation, length of stay, rates of ICU admission and mortality of the COVID groups. The Kruskal-Wallis test was used to analyze the other variables in the three study groups (IQR = interquartile range).

| Clinical characteristics | HEALTHY | COVID-19 | COVID-19 AKI | p-value |
| --- | --- | --- | --- | --- |
| Age (years, median and IQR) | 63 (18) | 64,5 (31) | 64 (35) | 0.816 |
| Gender (male) | 9 (60%) | 25 (69.4%) | 12 (70.6%) | 0.771 |
| Hypertension | 7 (46.7%) | 22 (61.1%) | 12 (70.6%) | 0.382 |
| Diabetes | 6 (40%) | 15 (41.7%) | 8 (47.1%) | 0.908 |
| Heart Failure | 1 (6.7%) | 1 (2.8%) | 2 (11.8%) | 0.426 |
| Coronary Artery Disease | 2 (13.3%) | 3 (8.3%) | 2 (11.8%) | 0.844 |
| Asthma or COPD | 1 (6.7%) | 2 (5.6%) | 1 (5.9%) | 0.988 |
| Obesity | 3 (20%) | 6 (16.7%) | 2 (11.8%) | 0.814 |
| Symptoms duration prior to admission (days, median and IQR) | - | 9.5 (14) | 7 (11) | 0.351 |
| Mechanical Ventilation | - | 2 (5.6%) | 8 (47.1%) | < 0.001 |
| Length of stay (days, median and IQ) | - | 13.5 (83) | 20 (49) | 0.005 |
| ICU admission | - | 16 (44.4%) | 16 (94.1%) | 0.001 |
| Outcome (deaths) | - | 4 (11.1%) | 11 (64.7%) | < 0.001 |

**Table 2.**
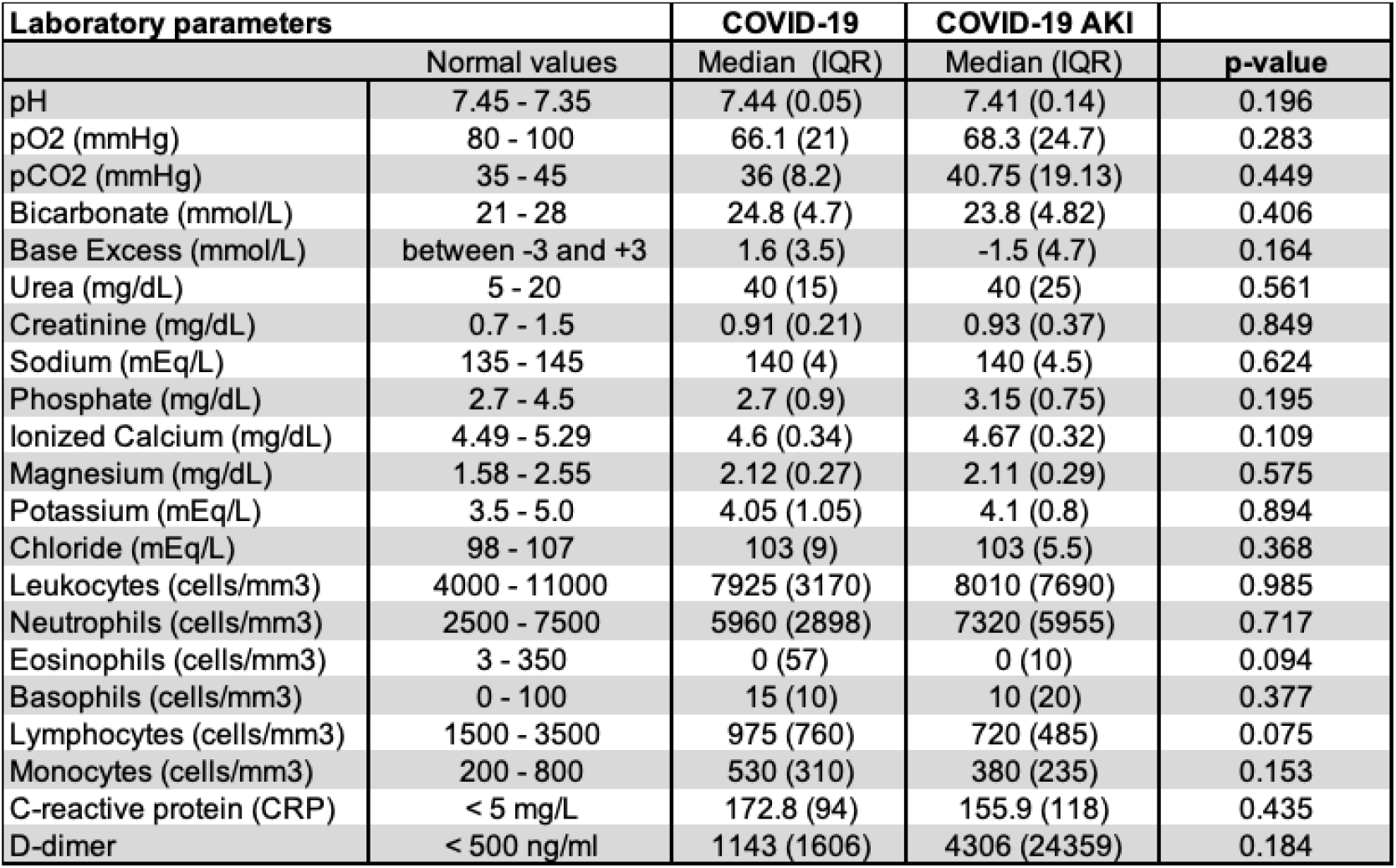
Laboratory parameters of the patients included in our study (COVID-19 group = 36 patients; COVID-19 AKI group = 17 patients). Mann-Whitney test was used to compare the COVID groups, normal values are exhibited only for reference (IQR = interquartile range).

### Sars-CoV-2 induced robust release of α-defensin 1, α-defensin 3 and β-defensin 3, but the LL-37 and β-defensin 1 secretion systems were not activated

Sars-CoV-2 infection induced potent release of α-defensin 1 (COVID-19 AKI versus healthy, p < 0.001; COVID-19 AKI versus COVID-19, p= 0.018) and α-defensin 3 (COVID-19 versus healthy, p < 0.001; COVID-19 AKI versus healthy, p < 0.001; COVID-19 AKI versus COVID-19, p = 0.05) in the plasma, especially in the COVID-19 AKI group (Figure 2A and 2B, respectively). β-defensin 3 levels also significantly increased following Sars-CoV-2 infection (COVID-19 versus healthy, p = 0.01; COVID-19 AKI versus healthy, p < 0.01) (Figure 2D). There was no difference, however, in β-defensin 1 plasma levels among the study groups (Figure 2C).

**Figure 2.**
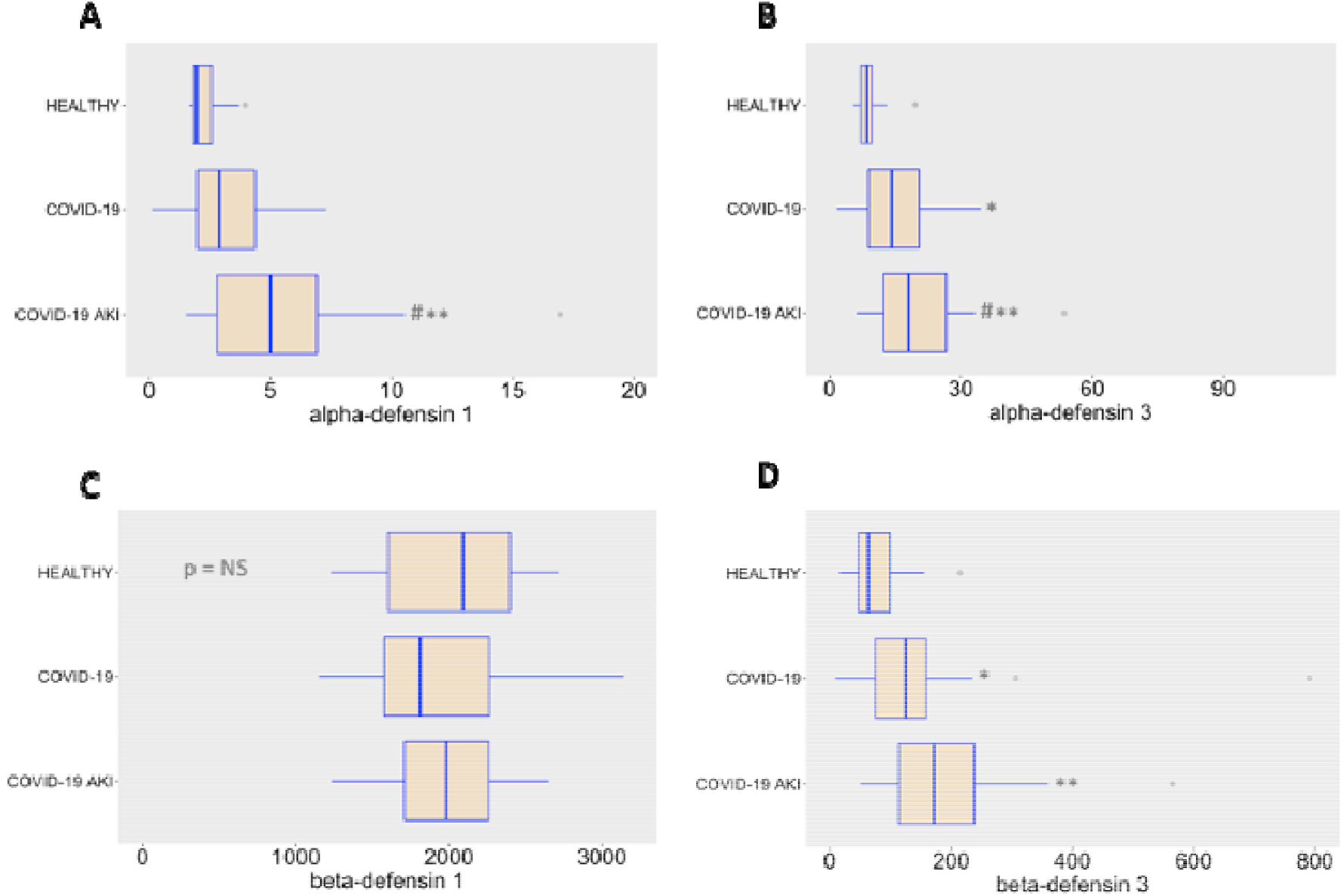
Plasma levels of α-defensin 1 (A), α-defensin 3 (B), β-defensin 1 (C) and β-defensin 3 (D) in the study groups. * indicates significant difference between the healthy and the COVID-19 groups. ** indicates significant difference between the healthy and the COVID-19 AKI groups. # indicates significant difference between the COVID and the COVID-19 AKI groups. Grey circles represent outliers. α-defensin 1 and α-defensin 3 values are in ng/ml. β-defensin 1 and and β-defensin 3 values are in pg/ml.

The LL-37 plasma levels found in our COVID-19 patients are very similar to the basal levels, putting in evidence that the LL-37 secretion system did not respond either (Figure 3). Important to cite, a previous study from our group showed inhibition of LL-37 gene expression during septic shock, in comparison with patients in severe sepsis, while LL-37 plasma levels in both severe sepsis and septic shock remained at similar levels than the healthy controls [24]. These results put in evidence that the LL-37 pathway doesn’t respond as expected in the presence of vigorous systemic inflammation. Since cathelicidins are able to induce both pro-or anti-inflammatory responses [15], it is difficult to conclude if this kind of inhibition is protective to the host or not. We cannot exclude, moreover, that it is a pathogen evasion strategy.

**Figure 3.**
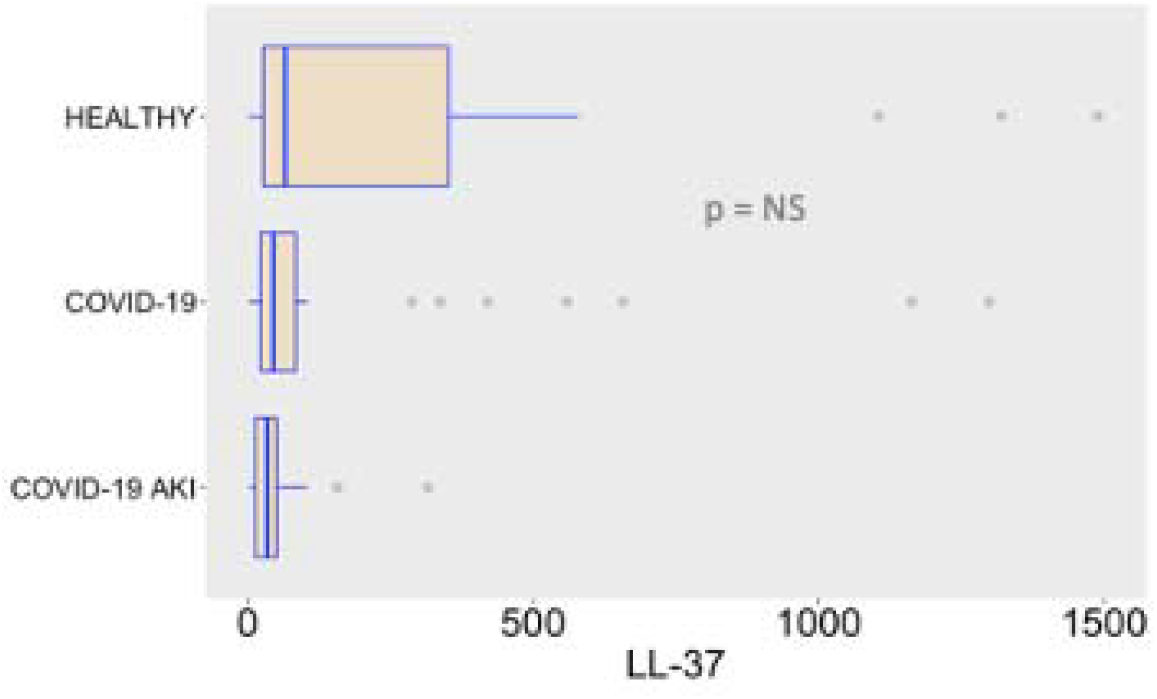
Plasma levels of LL-37 in the study groups. LL-37 values are in ng/ml.

### While IL-6, IL-10, IFN-γ and MCP-1 plasma levels significantly increased following Sars-CoV-2 infection, the TNF-α and IL-1β pathways remained undisturbed

COVID-19 induced potent secretion of IL-6 (COVID-19 versus healthy, p < 0.01; COVID-19 AKI versus healthy, p < 0.01; COVID-19 AKI versus COVID-19, p < 0.01), IL-10 (COVID-19 versus healthy, p < 0.01; COVID-19 AKI versus healthy, p < 0.01; COVID-19 AKI versus COVID-19, p < 0.01), IFN-γ (COVID-19 versus healthy, p < 0.01; COVID-19 AKI versus healthy, p < 0.01) and MCP-1 (COVID-19 versus healthy, p = 0.02; COVID-19 AKI versus healthy, p < 0.01; COVID-19 AKI versus COVID-19, p = 0.032) (Figures 4C, 4D, 5A and 5B, respectively). There was no significant secretion, however, of TNF-α and IL-1β in the COVID-19 groups, when compared with the healthy controls (Figures 4A and 4B).

**Figure 4.**
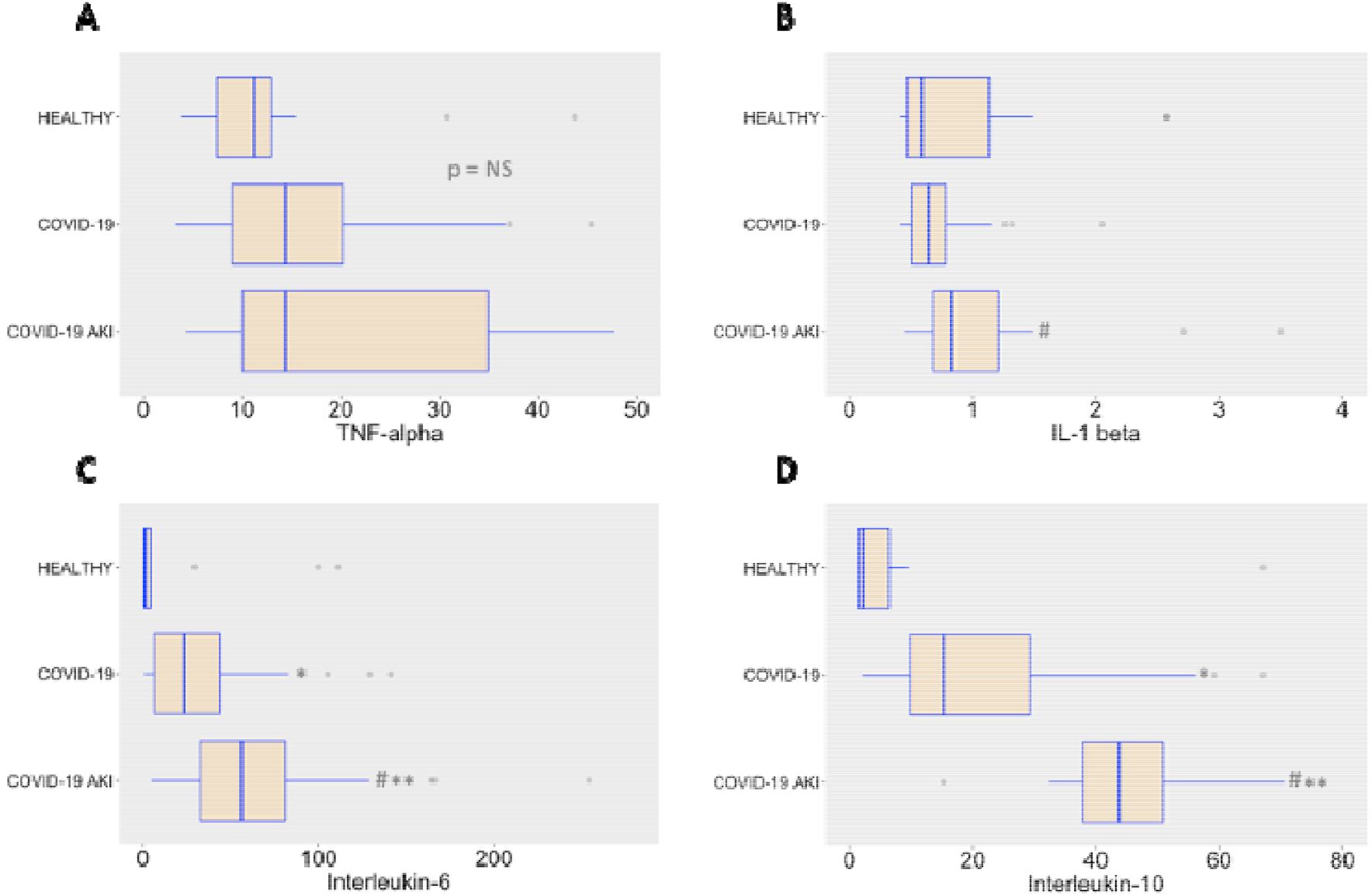
Plasma levels of TNF-α (A), IL-1β (B), IL-6 (C) and IL-10 (D) in the study groups. * Indicates significant difference between the healthy and the COVID-19 groups. ** indicates significant difference between the healthy and the COVID-19 AKI groups. # Indicates significant difference between the COVID and the COVID-19 AKI groups. Grey circles represent outliers. Cytokine values are in pg/ml.

**Figure 5.**
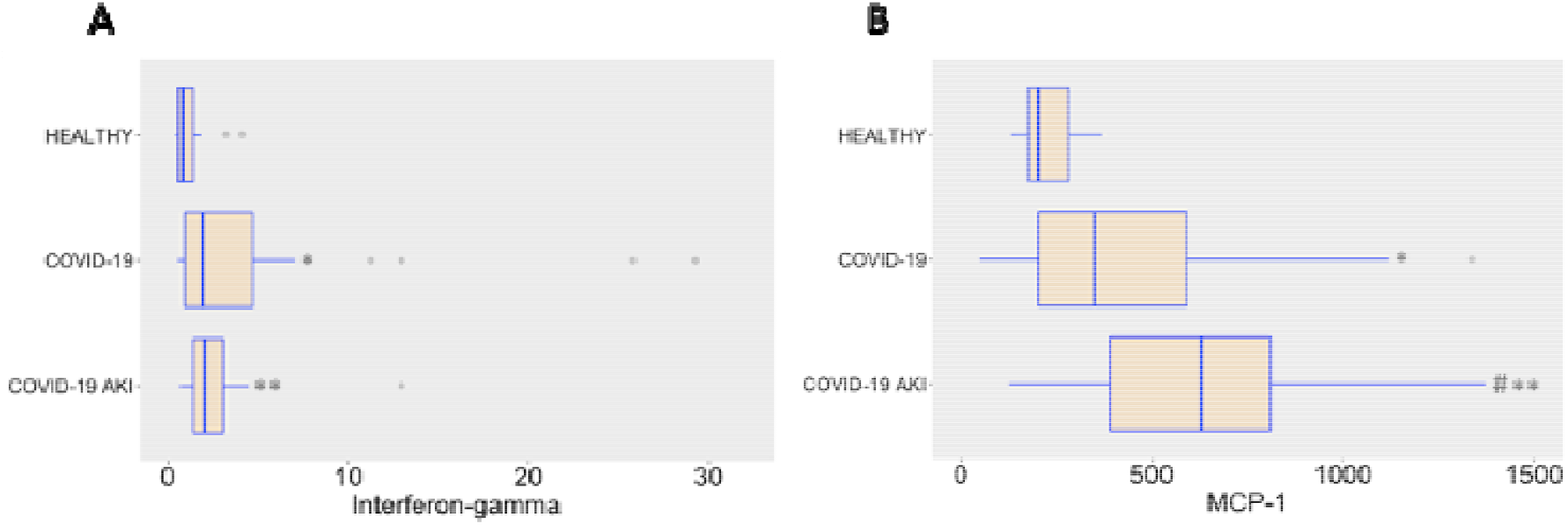
Plasma levels of IFN-γ (A) and MCP-1 in the study groups. * Indicates significant difference between the healthy and the COVID-19 groups. ** indicates significant difference between the healthy and the COVID-19 AKI groups. # Indicates significant difference between the COVID and the COVID-19 AKI groups. Grey circles represent outliers. Cytokine values are in pg/ml.

We believe that differences in TNF-α and IL-1β plasma levels could not be detected in the COVID-19 groups, in comparison with the healthy individuals, because the cytokine storm is milder in Sars-CoV-2 infections than in bacterial sepsis [25], being more difficult to detect. The other cytokines, however, clearly indicate that systemic inflammation is a hallmark of severe COVID-19 infections, occurs in association with the development of AKI and several antimicrobial peptides, such as α-defensin 1, α-defensin 3 and β-defensin 3 are also implicated in this mechanism of disease. Indeed, higher levels of IL-6 and IL-10 have been described as predictors of more severe disease in COVID-19 [26]. The role of defensins, however, cannot be oversimplified. Since defensins, in the same way as cathelicidins, can also exhibit both inflammatory and anti-inflammatory properties [27], further studies are necessary for a deeper comprehension of their activity, when Sars-CoV-2 spreads throughout the body. In what concern to cytokines, a study with patients undergoing cardiac surgery showed that both IL-6 and IL-10 are related to AKI. It also showed that IL-10 has a significant statistical correlation with levels of neutrophil gelatinase-associated lipocalin, a well-known biomarker of AKI [28]. In COVID-19 patients, elevated IL-10 is a strong predictor of severe disease, as shown by a recent study [29, 30]. The role of IL-10 in the pathophysiology of AKI is not clearly understood but it seems that it has a protective role, at least in some AKI experimental models such as ischemia-reperfusion related AKI [31]. In multiple experimental models of AKI, the role of IL-1β as a promoter of inflammation and as a marker of worse renal outcome are described [32]. In COVID-19 patients IL-1β has been perceived in bronchoalveolar lavage fluid and this cytokine seems to be related with the severe spectrum of this disease [33]. Despite that, Medeiros et. al. found a poor correlation between IL-1β levels and SARS-CoV2 viral load [34].

### IL-10 and the product IL-10 x IL-1B showed a good performance in ROC curve analysis and IL-10 showed a good correlation coefficient related to AKI

As shown in Figure 6, both IL-10 and the product IL-10 x IL-1β had a good performance in discriminate AKI from non-AKI patients. The area under the curve (AUC) for IL-10 was 0.86 (CI 0.77 – 0.94) and for the product IL-10 x IL-1β was 0.88 (CI 0.80 – 0.97), both with a p-value < 0,001. The optimum ROC curve point (defined by Youden’s index – sensitivity + specificity - 1) for IL-10 (cut-off 30.1 pg/mL) showed a sensitivity of 94% and a specificity of 75%, while the optimum ROC curve point for the product IL-10 x IL-1β (cut-off 24.48 pg2/mL2) resulted in a sensitivity of 88% and a specificity of 83%. Table 3 shows the AUC, sensitivity, specificity and cut-off of another cytokines and AMPs as well. Although a causal relationship isn’t possible to infer with our data, it’s of notice that IL-10 may be related to the pathophysiology of AKI in COVID patients (Pearson correlation coefficient between IL-10 and the Highest Creatinine, a surrogate of AKI, was 0.508, p-value < 0,001), a finding that contradicts the findings of another study with an experimental model of ischemia-reperfusion AKI in which IL-10 showed potential of protective effect [35].

**Table 3.** ROC curve analysis of selected cytokines and AMPs to differentiate COVID-19 with AKI from COVID-19 patients

| Biomarker | AUC | 95% CI | Cut-off | Sens | Spec | p-value |
| --- | --- | --- | --- | --- | --- | --- |
| IL-6 (pg/mL) | 0.76 | 0.65-0.87 | 30.4 | 82% | 64% | 0.002 |
| IL-10 (pg/mL) | 0.86 | 0.77-0.94 | 30.1 | 94% | 75% | <0.001 |
| IL-1B (pg/mL) | 0.73 | 0.61-0.85 | 0.6 | 88% | 44% | 0.008 |
| MCP-1 (pg/mL) | 0.68 | 0.56-0.81 | 386.0 | 76% | 58% | 0.031 |
| DEFA1 (ng/mL) | 0.70 | 0.57-0.83 | 3.33 | 71% | 61% | 0.018 |
| DEFA3 (ng/mL) | 0.67 | 0.53-0.80 | 14.16 | 71% | 50% | 0.052 |
| IL-10 x IL1B (pg<sup>2</sup>/mL<sup>2</sup>) | 0.88 | 0.80-0.97 | 24.484 | 88% | 83% | <0.001 |

**Figure 6.**
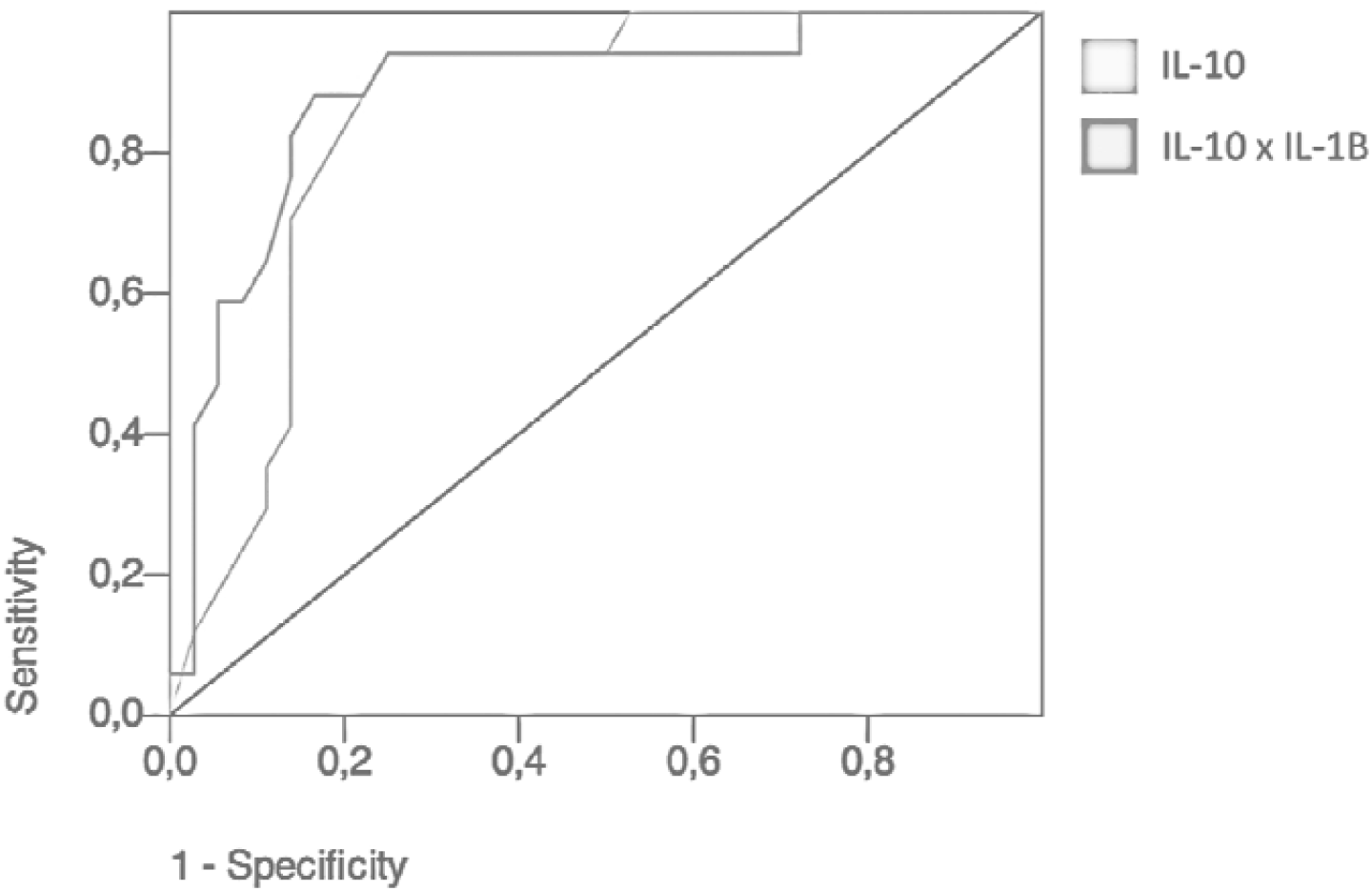
ROC Curve for IL-10 and the product IL-10 x IL-1β to differentiate COVID-19 with AKI from COVID-19 patients.

## Conclusion

The robust release of α-defensin 1 and α-defensin 3 in patients with COVID-19 and AKI in comparison with those with COVID-19 alone is of interest because it may be a potential pathway to reduce the development of AKI. Moreover, given our findings, the role of IL-10 in the pathophysiology of AKI must be further investigated so we have a better understanding of AKI in COVID-19.

In the last few years, the struggle to find better biomarkers of AKI other than creatinine resulted in the description of many biomarkers, like KIM-1, NGAL, TIMP-2 and IGFBP7. Although none of them are perfect, the product TIMP-2 x IGFBP7 has shown good performance in the Sapphire study, with an AUC of 0.80. In our study, the product IL-10 x IL-1β showed an AUC of 0.88 to discriminate AKI. Of notice, our blood samples were collected in the first 24h of hospitalization, before the detection of AKI, as defined according to creatinine criteria. Further studies are needed to define whether these biomarkers have such a good performance in a bigger and/or diverse population, so we can implement them in the clinical practice.

## Data Availability

All data produced in the present study are available upon reasonable request to the authors.

## Abbreviations Page

ACE2: Angiotensin-converting enzyme 2
AIDS: Acquired immunodeficiency syndrome
AKI: Acute Kidney Injury
AMPs: Antimicrobial peptides
COVID-19: Coronavirus Disease 2019
CRAMP: Cathelin-related antimicrobial peptide
HDP: Host defense peptides
HIV: Human immunodeficiency virus
ICU: Intensive care unit
IFN-γ: Interferon-γ
IL-1β: Interleukin-1β
IL-6: Interleukin-6
IL-10: Interleukin-10
IQR =: interquartile range
MCP-1: monocyte chemoattractant protein-1
RT-PCR: Real-time polymerase chain reaction
Sars-CoV-2: Respiratory syndrome coronavirus 2
TNF-α: Tumor necrosis factor-α

## Acknowledgements

FPS and HPS are supported by FAPESP, the São Paulo Research Foundation (grants # 2020/03905-8, 2016/14566-4 and 2020/04738-8).

FPS is supported by CNPq, the National Council for Scientific and Technological Development (grant # 303924/2018-7).

We would like to express our gratitude to the patients and their families.

## Authorship

FPS and HPS conceived the study. LFTS and HVB collected the samples. LFTS, HVB and DFB performed the experiments. LFTS and FPS performed the statistical analysis. FPS and HPS supervised the project. LFTS wrote the first draft. FPS wrote the final manuscript.

